# Genotype Distribution and Noninvasive Fibrosis Assessment in Chronic HCV - Insights from a Tertiary Center in Eastern India

**DOI:** 10.1101/2025.06.09.25329252

**Authors:** Jayeeta Bhowmick, Ranjit Das, Dipak Barik, Arka Bagchi, M Mugunthan, Sayantan Banerjee

**Author notes:** **Corresponding author:** Sayantan Banerjee.

## Abstract

**Introduction:** Hepatitis C virus (HCV) genotype distribution and fibrosis burden vary geographically and influence clinical management. Genotype 3 predominates in India and neighboring regions and is associated with steatosis and faster fibrosis progression. This study aims to characterize HCV genotypes, liver stiffness (FibroScan), and the comparative performance of non-invasive fibrosis markers (APRI, FIB-4) in patients from AIIMS Kalyani, West Bengal.

**Methods:** Between March 2023 and March 2025, a total of 1,105,120 patients attended the outpatient department and 33,287 were admitted to inpatient care at AIIMS Kalyani. Among them, 29,169 individuals were screened for anti-HCV antibodies. HCV seropositivity was detected in 140 patients (0.48%). Among these, 120 underwent HCV RNA PCR testing, of which 32 were confirmed RNA-positive and were enrolled for further evaluation in the Viral Hepatitis Treatment Clinic. Twenty seropositive patients were lost to follow-up. Detailed clinical, laboratory, and liver fibrosis assessments (FibroScan, APRI, FIB-4) were conducted in these 32 patients.

**Results:** Among 32 HCV RNA-positive patients, the mean age was 42.1 years, with 22 males (68.8%). Genotype 3 was predominant (43.8%), followed by genotype 1 (25%). Cirrhosis was detected in 15.6% (5/32) by FibroScan. FIB-4 >3.25 showed 80% sensitivity and 95.5% specificity (AUROC 0.96) for detecting cirrhosis, outperforming APRI >1.5, which had 40% sensitivity and 90.9% specificity (AUROC 0.840). These results support the FIB-4 as a reliable, low-cost alternative to FibroScan for assessing fibrosis in low-resource settings.

**Conclusion:** From a large population screened over two years, only a small fraction demonstrated active HCV infection requiring treatment. Genotype 3 remains the most prevalent in Eastern India, associated with steatosis and progressive fibrosis. FIB-4 provides superior non-invasive prediction of cirrhosis compared to APRI, supporting its use in resource-limited settings.

## Introduction

Hepatitis C virus (HCV) infection is a major cause of chronic liver disease globally, affecting an estimated 58-71 million people worldwide [1]. HCV shows extensive genetic heterogeneity with at least seven genotypes and 67+ subtypes identified. The distribution of HCV genotypes varies markedly by geography [1,2]. Globally, genotype 1 is most prevalent (∼46% of cases), followed by genotype 3 (∼22%), with genotypes 2, 4, and others less common [1]. In South Asia, and particularly in India and its neighboring countries, genotype 3 predominates in most regions [3,4]. Studies from Northern, Eastern, and Western India consistently report genotype 3 as the most common, whereas Southern India shows a more equal mix of genotypes 1 and 3 [3,5]. For example, a large multicenter Indian survey found genotype 3 in 54% of chronic HCV cases nationally (and 69% in Eastern India), with genotype 1 comprising ∼24% and genotype 4 around 6% of cases; in 16% of patients, genotype could not be determined by standard assays [6]. Similarly, in Bangladesh, genotype 3 is the predominant strain (50-89% of infections) followed by genotype 1, with other genotypes (2, 4, 6) being rare; about 5-10% of isolates may remain untyped by routine PCR methods [7,8].

Adjacent regions of Southeast Asia also have distinctive patterns: genotype 3 is common in countries like Thailand and Malaysia, genotype 1 dominates in some urbanized areas (e.g., Singapore), and genotype 6 is highly prevalent in parts of Myanmar, Laos, and Vietnam [9]. These regional differences in genotype distribution have clinical significance. Genotype determination is important for treatment decisions and prognostication - for instance, genotype 3 has historically shown somewhat lower response rates to interferon-based therapy and is now known to confer a higher risk of liver disease complications even after antiviral cure [10,11].

Besides influencing therapy, HCV genotype appears to impact disease phenotype. Infection with genotype 3 has been strongly linked to hepatic steatosis (fatty liver). Early studies demonstrated that genotype 3 induces a direct cytopathic effect leading to fat accumulation in hepatocytes, independent of metabolic factors [12]. In a landmark study by Rubbia-Brandt et al., patients with genotype 3 had significantly higher grades of steatosis, and successful antiviral treatment led to the disappearance of fatty changes, implicating the virus in causing steatosis [12]. Genotype 3 infection is also associated with accelerated fibrosis progression in many cases. Recent evidence indicates that even after achieving sustained virological response (SVR) with direct-acting antivirals, patients with genotype 3 remain at elevated risk of developing cirrhosis and hepatocellular carcinoma (HCC) compared to non-GT3 patients [11]. This contrasts with genotypes 1 or 2, where liver outcomes post-cure are generally more favourable. Thus, understanding genotype distribution in a population is vital for anticipating disease burden - genotype 3 predominance could portend more cases of steatohepatitis, advanced fibrosis, and HCC down the line [4,10,11].

Eastern India, including the state of West Bengal, lies at an epidemiologic crossroads adjoining Bangladesh and other high-prevalence areas. Prior studies in West Bengal’s general population found a relatively low HCV seroprevalence (∼0.9%), but an overwhelming dominance of genotype 3 among those infected (over 85-90% in some surveys) [3]. However, many of these data are from early 2000s or special groups (e.g., multi-transfused thalassemia patients) [3,4,13]. With the advent of highly effective antiviral therapy, updated local data on genotype patterns and liver disease status are needed to inform public health and clinical strategies. In particular, non-invasive assessment of liver fibrosis, such as via Transient Elastography (FibroScan®), has become routine in HCV management and can stratify patients by fibrosis stage without biopsy. Large Indian studies have validated FibroScan’s accuracy in chronic liver diseases [14], but there is limited published data correlating FibroScan findings with genotypes or other clinical parameters in our region.

This study was undertaken to characterize the genotypic distribution of HCV and associated clinical features in a group of patients presenting to a tertiary care hospital from Eastern India (Kalyani, West Bengal). We additionally evaluated liver stiffness and steatosis quantitatively using FibroScan, and explored correlations between genotype, liver stiffness, and biochemical markers. We also performed an exploratory cluster analysis of patient data to identify any natural groupings (for example, clusters of patients with particular genotype and liver stiffness profiles). By comparing our findings with those from neighboring regions (e.g., Bangladesh, Northeast India, Southeast Asia), we aim to place the HCV epidemic in Eastern India in context. Ultimately, this information can help optimize screening and management - for instance, highlighting the need to screen for steatosis in genotype 3 patients, or the importance of sensitive genotyping methods to detect uncommon strains in this region.

## Materials And Methods Study

### Design and Patients

We conducted a cross-sectional analysis of patients diagnosed with chronic hepatitis C infection at AIIMS Kalyani, a tertiary care teaching hospital in West Bengal, India. The study period spanned from March 2023 to March 2025. During this time, 1,105,120 patients attended the outpatient department and 33,287 were admitted. A total of 29,169 patients were tested for anti-HCV antibodies, of whom 140 (0.48%) tested positive. HCV RNA PCR testing was performed in 120 seropositive individuals, and 32 patients were confirmed to have detectable HCV RNA, thus forming the basis of our study. Patients were eligible if aged ≥3 years and untreated for HCV. Those with prior antiviral therapy or co-infection with hepatitis B virus or HIV were excluded. Informed consent was obtained from all adult patients and from guardians in the case of pediatric patients. The study protocol was approved by the Institutional Ethics Committee of AIIMS Kalyani.

### Data Collection

For each participant, demographic data (age, sex), relevant medical history, and laboratory investigations were recorded. These included liver function tests (bilirubin, ALT, AST, alkaline phosphatase, total protein, albumin, globulin), HCV serology, RNA viral load, and genotype. Anti-HCV antibody detection was performed using a third-generation ELISA. Quantification of HCV RNA was conducted using the COBAS TaqMan real-time PCR platform (Roche Diagnostics), with results expressed in IU/mL.

### Genotyping

HCV genotyping was performed using the TruPCR^®^ HCV Genotyping Kit (Cat No. 3B296, 3B BlackBio Biotech India Ltd., Bhopal, India), a commercial multiplex RT-PCR assay targeting the 5’ untranslated region (5’UTR) and core regions of the HCV genome. The kit is designed to identify common genotypes 1a, 1b, 2, 3, 4, 5a, and 6 via genotype-specific probes. The assay has demonstrated robust concordance with sequencing-based genotyping in South Asian populations and complies with IVD quality certification [15]. In our study, genotyping was successfully performed in most RNA-positive samples; no untypable strains were recorded in the final dataset. Missing genotype data in a few patients were due to logistical limitations or attrition.

### FibroScan Assessment

Transient elastography was performed using FibroScan^®^ 402 (Echosens, Paris). The procedure was carried out on fasting patients by a trained operator with 10 successful acquisitions, and the median value was recorded. Liver stiffness values were categorized as F0-F1 (<7.1 kPa), F2 (7.1-9.4), F3 (9.5-12.4), and F4 (≥12.5). CAP values above 248 dB/m were considered indicative of moderate-to-severe steatosis [16].

### Statistical Analysis

Data were analyzed using SPSS v25 and Python (SciPy). Descriptive statistics (median, IQR; percentages) were used. Between-group comparisons (e.g., genotype groups) employed Fisher’s exact or Mann-Whitney U test. Correlation between continuous variables (ALT, AST, CAP, viral load, and FibroScan kPa) was tested using Spearman’s rank correlation. ROC curves were generated to assess APRI and FIB-4 scores’ predictive value for cirrhosis (FibroScan ≥12.5 kPa). Cut-offs were optimized using Youden’s index. Hierarchical and k-means clustering (k=3) were applied to FibroScan, CAP, ALT, AST, and viral load for pattern recognition.

## Results

### Demographic and Clinical Profile

Between March 2023 and March 2025, a total of 1,105,120 patients attended outpatient services, and 33,287 were admitted at AIIMS Kalyani, a tertiary care center in Eastern India. During this period, 29,169 patients underwent anti-HCV serological testing, of whom 140 (0.48%, N=29,169) tested positive. Among these, 120 patients underwent HCV RNA PCR testing, and 32 patients were confirmed to have detectable HCV RNA, thereby forming the study cohort for the final analysis. This stepwise diagnostic cascade illustrates the burden of infection and the systematic filtering required to identify active cases. A total of 32 patients with detectable HCV RNA were included in the study. Of these, 22 (68.8%) were male. The mean age was 42.1 years (Standard Deviation ± 18.9), with a median age of 41 years (interquartile range 32-60) (Table *1*).

**TABLE 1:** Demographic and Clinical Profile of HCV RNA-Positive Patients (N=32) Data represented as N (%) for categorical variables and median (IQR) for continuous variables. Age range provided as minimum to maximum values in completed Years.

| Variable | Value |
| --- | --- |
| Total patients with HCV RNA detected | 32 (100%) |
| Male sex | 22 (68.8%) |
| Mean age (years) ± SD | 42.1 ± 18.9 |
| Median age (IQR) | 41 (32–60) |
| Pediatric/Adolescent patients | 3 (9.4%) |

### Laboratory Parameters and Liver Function Tests

Baseline liver function tests revealed elevated ALT levels (>40 U/L) in 12 (37.5%) patients and elevated AST (>40 U/L) in nine (28.1%) patients. Serum bilirubin was elevated (>2 mg/dL) in five (15.6%) patients, while albumin levels were below normal (<3.5 g/dL) in three (9.4%) patients, indicating impaired liver function. Most patients (90.6%) had preserved synthetic liver function, with serum albumin ≥3.5 g/dL. Notably, patients with advanced fibrosis frequently presented only mildly elevated or normal transaminase levels, highlighting the limitations of ALT and AST alone in accurately predicting fibrosis severity (Table *2*).

**TABLE 2:** Laboratory Parameters and Liver Function Tests among RNA-positive HCV Patients (N=32). Data expressed as N (%) indicating the proportion of patients with elevated or reduced parameters. (ALT - alanine aminotransferase AST – aspartate aminotransferase).

| Parameter | Number Elevated | Percentage (%) |
| --- | --- | --- |
| ALT (>40 U/L) | 12 | 37.5 |
| AST (>40 U/L) | 9 | 28.1 |
| Bilirubin (>2 mg/dL) | 5 | 15.6 |
| Albumin (<3.5 g/dL) | 3 | 9.4 |

### HCV Genotype Distribution

Genotype testing was performed using the TruPCR^®^ HCV Genotyping Kit (3B BlackBio Biotech), with valid results in 22 out of 32 patients. Genotype 3 was the most prevalent, detected in 14 patients (43.8%; N=32), followed by genotype 1 in 8 patients (25% of N=32). Ten patients (31.3%; N=32) lacked genotype data due to logistical issues or follow-up attrition (Table *3*). Among genotyped cases, 63.6% (14 out of 22) were genotype 3 and 36.4% (8 out of 22) genotype 1. Genotype 1 infections were more common among older adults (mean age 52 years vs. 43 years in genotype 3), suggesting legacy infections possibly related to transfusion practices before the implementation of strict blood screening protocols.

**TABLE 3:** HCV Genotype Distribution Among RNA-Positive Patients (N=32) Data represented as N (%) of total RNA-positive patients. Successful genotyping was performed in 22 cases, with the remaining 10 cases (31.2%) categorised as Nontyped (due to logistic constraints).

| Genotype | Number | Percentage (%) |
| --- | --- | --- |
| 3 | 14 | 43.8 |
| 1 | 8 | 25.0 |
| Nontyped (due to logistic constraints) | 10 | 31.2 |

### Viral Load Trends

Among the 22 genotyped patients, genotype 3 cases (n = 14) had a median viral load of 5.67 × 10^5^ IU/mL (IQR: 1.74 × 10^5^ - 2.87 × 10^6^ IU/mL) and a mean of 2.02 × 10^6^ IU/mL, with a range of 4.43 × 10^4^ to 8.48 × 10^6^ IU/mL. Genotype 1 cases (n=8) had a median viral load of 5.03×10^5^ IU/mL (IQR: 6.55×10^5^ -3.14×10^6^ IU/mL), and a mean of 2.05×10^6^ IU/mL, with values ranging from 6.04×10^4^ to 7.85×10^6^ IU/mL. No statistically significant difference in median viral load was observed between genotypes 1 and 3 (p=0.45; Mann-Whitney U test). Viral load did not correlate significantly with fibrosis stage (Spearman’s rho <0.1), emphasizing that viral load alone is a poor surrogate for fibrosis assessment (Table *4*).

**TABLE 4:**
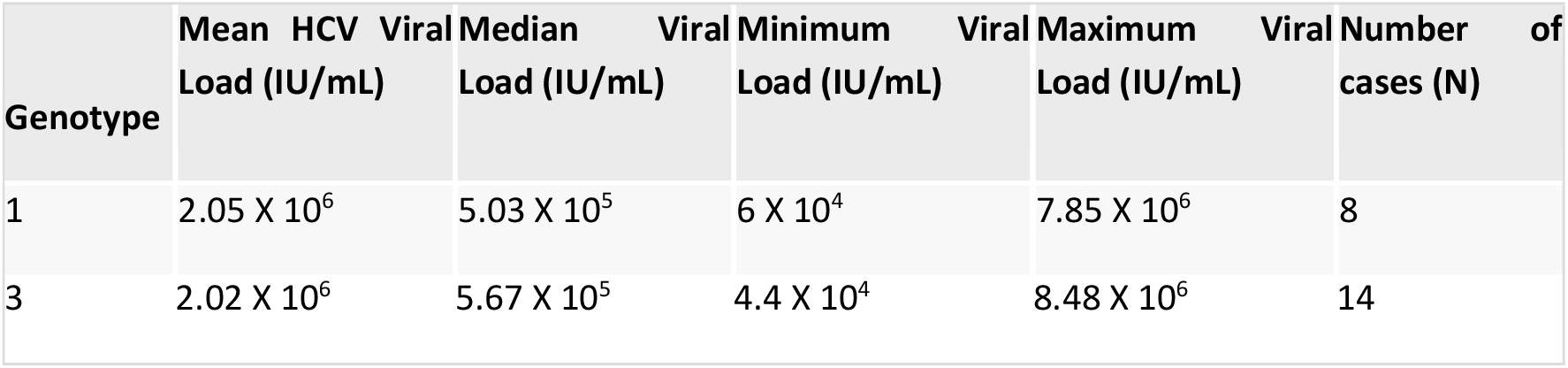
Viral Load Distribution According to HCV Genotype (IU/mL) in Genotyped Patients (N=22). Data reported as median (IQR), mean, minimum, and maximum. Statistical test: Mann– Whitney U test. Viral load expressed in IU/mL. Only genotyped cases are included.

### Fibrosis and Steatosis (FibroScan, APRI scoring and FIB-4 scoring)

In this real-world cohort of 32 untreated chronic hepatitis C patients attending the Viral Hepatitis Clinic at AIIMS Kalyani, Eastern India, liver fibrosis severity was assessed by FibroScan and compared with APRI and FIB-4 indices.

Transient elastography (FibroScan) data were available for 23 of the 32 HCV RNA-positive patients. Among these, liver stiffness measurements indicated F0-F1 (no or mild fibrosis; <7.1 kPa) in 12 patients (52.2%), F2 (moderate fibrosis; 7.1-9.4 kPa) in 4 patients (17.4%), F3 (advanced fibrosis; 9.5-12.4 kPa) in 2 patients (8.7%), and F4 (cirrhosis; ≥12.5 kPa) in 5 patients (21.7%). Thus, 30.4% (7 out of 23) of the patients had advanced fibrosis (F3 or F4), despite being mostly treatment-naïve and relatively young.

Steatosis assessment using the controlled attenuation parameter (CAP) revealed that 2 out of 23 patients (8.7%) had moderate-to-severe hepatic steatosis, defined as CAP ≥250 dB/m. The fibrosis stages used standard thresholds, and non-invasive fibrosis indices-APRI and FIB-4-were stratified by standard cut-offs: APRI (<0.5, 0.5-1.5, >1.5) and FIB-4 (<1.45, 1.45-3.25, >3.25).

We evaluated the diagnostic accuracy of APRI and FIB-4 scores against FibroScan as the reference standard for cirrhosis (defined as ≥12.5 kPa). FibroScan staging was categorized as F0-F1 (<7.1 kPa), F2 (7.1-9.4), F3 (9.5-12.4), and F4 (≥12.5). APRI and FIB-4 were stratified using standard thresholds: APRI (<0.5, 0.5-1.0, 1.0-1.5, >1.5) and FIB-4 (<1.45, 1.45-3.25, >3.25). Among the five patients with cirrhosis, an APRI >1.5 correctly identified 2 cases (sensitivity 40%), while FIB-4 >3.25 identified 4 of 5 cases (sensitivity 80%). Specificity among non-cirrhotic cases was 90.9% for APRI >1.5 and 95.5% for FIB-4 >3.25, underscoring the higher discriminative power of FIB-4 for cirrhosis detection.

Using APRI <0.5 as a rule-out threshold yielded a sensitivity of 60% and specificity of 81.8%, with a negative predictive value (NPV) of 90% and a positive predictive value (PPV) of 43%. Although APRI <0.5 provides reasonable confidence in excluding cirrhosis, it may still miss a significant fraction of true cirrhotic cases. Conversely, APRI >1.5 showed high specificity (90.9%) but modest sensitivity (40%) and PPV (50%), limiting its utility as a rule-in tool in isolation.

FIB-4 <1.45 demonstrated a sensitivity and NPV of 100%, making it an excellent tool to exclude cirrhosis in low-resource settings. Its specificity was 59.1% and PPV was 38.5%. At the higher threshold of >3.25, FIB-4 achieved superior diagnostic metrics: 80% sensitivity, 95.5% specificity, 80% PPV, and 95.5% NPV. These findings support FIB-4 as both a reliable rule-in and rule-out noninvasive index in this cohort (Table *5*).

**TABLE 5:** Comparative Performance of APRI and FIB-4 Scores against FibroScan for Detection of Cirrhosis (F4, ≥12.5 kPa). Data derived from a cohort of 23 patients with available FibroScan results. Cirrhosis was defined as liver stiffness measurement ≥12.5 kPa. Sensitivity, specificity, positive predictive value (PPV), and negative predictive value (NPV) were calculated using 2×2 contingency tables. AUROC (Area Under the Receiver Operating Characteristic Curve) represents the overall diagnostic performance based on ROC analysis. FIB-4 showed superior performance to APRI in detecting cirrhosis, as confirmed by DeLong’s test (p=0.04). Significance set at p<0.05. APRI - Aspartate Aminotransferase-to-Platelet Ratio Index FIB-4 - Fibrosis Index Based on 4 Factors PPV - Positive Predictive Value NPV - Negative Predictive Value AUROC - Area under the Receiver Operating Characteristic Curve

|  |  |  |  |  |  |  |
| --- | --- | --- | --- | --- | --- | --- |
| APRI | <0.5 | 60% | 81.8% | 43% | 90% | 0.84 |
| APRI | >1.5 | 40% | 90.9% | 50% | 87% | 0.84 |
| FIB-4 | <1.45 | 100% | 59.1% | 38.5% | 100% | 0.96 |
| FIB-4 | >3.25 | 80% | 95.5% | 80% | 95.5% | 0.96 |

Receiver Operating Characteristic (ROC) analysis further confirmed the superior performance of FIB-4. The Area Under the ROC Curve (AUROC) for FIB-4 was 0.96, compared to 0.84 for APRI. DeLong’s test for comparison of AUROCs yielded a statistically significant difference (p=0.04), favoring FIB-4 as the more robust fibrosis assessment tool. These findings reinforce the role of FIB-4 in routine clinical practice, especially where access to transient elastography remains limited.

## Discussion

This study presents a real-world, data-driven profile of chronic HCV infection in Eastern India, highlighting the enduring predominance of genotype 3 and the silent progression of fibrosis even in young patients.

These findings not only corroborate earlier studies from the region [3-7] but also offer comparative insights into noninvasive fibrosis staging tools validated in South Asian and global contexts.

### Epidemiology and Genotype Profile

The predominance of genotype 3 (43.8%) aligns with the genotype distribution reported by Chowdhury et al. in West Bengal [3], Shah et al. across India [6], Dutta et al. in multi-transfused populations [4] and that of Bangladesh [7]. Genotype 3 has been historically associated with transfusion-related infections, and our cohort’s pediatric cases with thalassemia reinforce this link [13]. The absence of genotypes 2, 4, 5 or 6 is not surprising, aligns with regional epidemiological trends, given their low prevalence in this region [5,7,17].

Interestingly, genotype 1 infections in our cohort were associated with older age, supporting the hypothesis that these may represent legacy cases predating stringent blood safety protocols. The implications are significant: as pan-genotypic DAAs become more accessible, the precise genotype may no longer guide therapy selection, but its association with disease phenotype remains clinically relevant.

### Liver Enzymes, Viral Load, and Disease Severity

Liver enzyme levels (ALT, AST) did not reliably predict fibrosis stage, consistent to earlier reports [12], with nearly half of patients with F3-F4 fibrosis showing only mild enzyme elevation. This observation aligns with Singh et al. [14] and emphasizes the limitations of biochemical markers alone for staging. Similarly, viral load had no correlation with fibrosis, similar to previous findings [6,7,10], reinforcing existing evidence that HCV RNA levels are not reflective of liver injury [4,8,14].

### Fibrosis Assessment: APRI, FIB-4, and FibroScan

A central component of our analysis was the evaluation of noninvasive fibrosis indices-APRI and FIB-4-against FibroScan.

In this cohort of chronic HCV patients, we evaluated the utility of APRI and FIB-4-two widely accessible non-invasive indices-for diagnosing or excluding cirrhosis, using transient elastography (FibroScan) as the reference standard. Among the 23 patients with available FibroScan data, five had cirrhosis (F4), and seven had advanced fibrosis (F3/F4), highlighting a significant burden of liver damage even in a relatively young and treatment-naïve population.

FIB-4 demonstrated superior diagnostic accuracy across all metrics. At a cut-off of <1.45, FIB-4 had 100% sensitivity and negative predictive value (NPV), making it highly effective for excluding cirrhosis. However, its specificity (59.1%) and positive predictive value (PPV, 38.5%) were modest, indicating limited rule-in utility at this threshold. In contrast, FIB-4 >3.25 offered both high sensitivity (80%) and specificity (95.5%), along with balanced PPV and NPV (both 80% and 95.5%, respectively), supporting its use as a robust rule-in test.

APRI, though less powerful, still showed utility. At <0.5, it yielded 60% sensitivity and 81.8% specificity, with an NPV of 90%, making it moderately reliable for excluding cirrhosis in resource-limited settings. APRI >1.5 demonstrated high specificity (90.9%) but low sensitivity (40%) and PPV (50%), limiting its use as a confirmatory tool.

Receiver Operating Characteristic (ROC) analysis further confirmed these trends. FIB-4 exhibited a superior Area Under the ROC Curve (AUROC) of 0.96, compared to APRI’s AUROC of 0.84. The DeLong’s test for comparing AUROCs yielded a statistically significant result (p=0.04), establishing FIB-4’s greater discriminatory power. These findings align with prior studies and reinforce FIB-4’s role as a more reliable index for staging fibrosis in low-resource settings where FibroScan may not be routinely available.

Overall, while both indices offer accessible alternatives to elastography, FIB-4 emerges as the preferred choice due to its superior accuracy for both ruling in and ruling out cirrhosis. Given the high burden of undiagnosed liver fibrosis and limited access to advanced imaging in the Global South, incorporating FIB-4 into clinical algorithms could significantly improve early detection and risk stratification in chronic HCV management. These results closely match the performance reported in Indian studies by Rungta et al. [18] and Kujur et al. [19], and global meta-analyses by Lin et al. and El-Kassas et al. [20,21]. They validate the use of these indices as first-line staging tools in resource-limited settings, where FibroScan is unavailable or inaccessible.

### Steatosis, Clustering, and Genotype 3 Pathogenesis

The steatogenic effect of genotype 3 was evident from our CAP values and clustering analysis. Grouping patients by fibrosis and steatosis levels showed that those with high fibrosis and high CAP were predominantly genotype 3. The association of steatosis with genotype 3 was evident, as was described before [2,5,10]. The finding echoes Rubbia-Brandt’s landmark description of genotype 3’s cytopathic steatosis [12] and newer findings linking genotype 3 to worse outcomes post-SVR [11].

**TABLE 6:** Comparison of our data with relevant previous studies between the AUROC values of APRI and FIB-4 with FibroScan or Biopsy as Reference Gold Standard. APRI - Aspartate Aminotransferase-to-Platelet Ratio Index FIB-4 - Fibrosis Index Based on 4 Factors PPV - Positive Predictive Value NPV - Negative Predictive Value AUROC - Area under the Receiver Operating Characteristic Curve

| Study | Country | Reference Std | APRI AUROC | FIB-4 AUROC | Remarks |
| --- | --- | --- | --- | --- | --- |
| Our Study (2025) | India | FibroScan | 0.844 | 0.941 | FIB-4 >3.25 highly specific; APRI >1.5 91% specificity |
| Rungta et al., 2021 [18] | India | FibroScan | 0.83 | 0.88 | FIB-4 >2.25 optimal for cirrhosis |
| Ishtiaq et al., 2021 [22] | Pakistan | FibroScan | ~0.70 | ~0.80 | FIB-4 better in indeterminate APRI |
| Ahsan et al., 2025 [23] | Pakistan | FibroScan | 0.866 | 0.994 | FIB-4 optimal ~2.44 for cirrhosis |
| Lin et al., 2011 [20] | Global meta | Biopsy | 0.77-0.80 | - | APRI validated vs METAVIR |
| El-Kassas et al., 2024 [21] | Egypt | Biopsy | ~0.79 | 0.82 | FIB-4 optimal ~2.2 for cirrhosis |

Our cluster analysis yielded three phenotypic groups: (i) High fibrosis and steatosis (mostly genotype 3), (ii) Low fibrosis and mild steatosis (younger, treatment-naïve) and (iii) Intermediate fibrosis without steatosis (older patients, likely genotype 1). Such phenotyping can aid targeted surveillance and treatment prioritization.

## Conclusions

This hospital-based study from Eastern India reinforces several important epidemiological and clinical observations regarding chronic hepatitis C. Genotype 3 remains the predominant strain in this region followed by genotype 1. These findings are consistent with prior studies from West Bengal and neighboring countries. Notably, genotype 1 infections were more common in older adults, suggesting legacy transmission through transfusion or dialysis prior to robust screening protocols.

Despite a relatively young cohort (median age 41 years), nearly one-third of patients exhibited advanced fibrosis or cirrhosis by FibroScan, underscoring the silent but progressive nature of HCV. Our study highlights the inadequacy of relying on liver enzyme elevations to predict fibrosis, as many patients with advanced liver disease had only mild biochemical abnormalities.

Importantly, noninvasive scores-FIB-4 and APRI-demonstrated strong concordance with FibroScan. FIB-4 >3.25 identified 100% of cirrhotic cases, and APRI >1.5 identified 60%, with both indices showing over 85% specificity. These findings support the adoption of tiered fibrosis staging strategies, where high-cost tests like FibroScan are reserved for indeterminate or high-risk noninvasive profiles.

In resource-limited settings across the Global South, this approach could significantly reduce dependence on expensive infrastructure while enabling timely care. We advocate the integration of validated APRI and FIB-4 thresholds into national HCV management protocols to improve accessibility, equity, and efficiency in the path toward hepatitis C elimination.

## Data Availability

All data produced in the present study are available upon reasonable request to the corresponding authors

## Acknowledgements

We acknowledge the VRDL Scheme of Department of Health Research, Govt. of India for infrastructural and financial support. We acknowledge the contribution of Mr. Kallal Biswas for the entry of data and statistical analysis, Mr. Aranyajit Samanta and Mr. Nur Habib Miah (Laboratory Technicians of AIIMS Kalyani) for the molecular analysis of HCV positive samples, Mrs. Surbhi Raykwar, Ms. Suchitra Mahato and Ms. Ananta Pramanik (Nursing Officer of AIIMS Kalyani) for the record maintenance, management and counselling of all Viral Hepatitis Clinic patients.

